# Genomic surveillance of SARS-CoV-2 in Thailand reveals mixed imported populations, a local lineage expansion and a virus with truncated ORF7a

**DOI:** 10.1101/2020.05.22.20108498

**Authors:** Elizabeth M Batty, Theerarat Kochakarn, Bhakbhoom Panthan, Krittikorn Kümpornsin, Poramate Jiaranai, Arporn Wangwiwatsin, Namfon Kotanan, Peera Jaruampornpan, Treewat Watthanachockchai, Kingkan Rakmanee, Insee Sensorn, Somnuek Sungkanuparph, Ekawat Pasomsub, Thanat Chookajorn, Wasun Chantratita

**Affiliations:** COVID-19 Network Investigations Alliance (CONI), Bangkok, Thailand.; Mahidol-Oxford Tropical Medicine Research Unit, Faculty of Tropical Medicine, Mahidol University, Bangkok, Thailand.; Centre for Tropical Medicine and Global Health, Nuffield Department of Medicine, University of Oxford, Oxford, United Kingdom.; Genomics and Evolutionary Medicine Unit (GEM), Center of Excellence in Malaria Research, Faculty of Tropical Medicine, Mahidol University, Bangkok, Thailand.; Faculty of Medicine Ramathibodi Hospital, Mahidol University, Bangkok, Thailand.; Center for Medical Genomics, Faculty of Medicine Ramathibodi Hospital, Mahidol University, Bangkok, Thailand.; Wellcome Sanger Institute, Hinxton, Cambridgeshire, United Kingdom.; Department of Biology, Faculty of Science, Khon Kaen University, Khon Kaen, Thailand.; Virology and Cell Technology Research Team, National Center for Genetic Engineering and Biotechnology (BIOTEC), National Science and Technology Development Agency (NSTDA), Pathum Thani, Thailand.; Division of Virology, Department of Pathology, Faculty of Medicine Ramathibodi Hospital, Mahidol University, Bangkok, Thailand.; Chakri Naruebodindra Medical Institute, Faculty of Medicine Ramathibodi Hospital, Mahidol University, Samut Prakan, Thailand.

**Keywords:** COVID-19, Genomic Surveillance, SARS-CoV-2, Thailand

## Abstract

Coronavirus Disease 2019 (COVID-19) is a global public health threat. Genomic surveillance of SARS-CoV-2 was implemented during March 2020 at a major diagnostic hub in Bangkok, Thailand. Several virus lineages supposedly originated in many countries were found, and a Thai-specific lineage, designated A/Thai-1, has expanded to be predominant in Thailand. A virus sample in the SARS-CoV-2 A/Thai-1 lineage contains a frame-shift deletion at ORF7a, encoding a putative host antagonizing factor of the virus.

## Introduction

Coronavirus Disease 2019 (COVID-19) has reached the status of global pandemic. Genomic surveillance of its etiological virus, SARS-CoV-2, plays an important role in epidemiological investigations and transmission control strategies [1]. Genetic variation data of the virus could reveal transmission chains between infected individuals and could even map the connection between outbreak cohorts. Thailand has suffered from the spread of COVID-19 with the total number of confirmed cases over 3,000 and with more than 120,000 individuals screened as of May 2020. Since January 2020, when both imported and locally-transmitted COVID-19 cases were reported in Thailand, the country has implemented several measures to combat COVID-19 at a national scale [2-4].

Genomic surveillance could be a powerful tool in the implementation of the national COVID-19 control strategy in Thailand. ARTIC multiplex tiling PCR allows whole-genome sequencing with minuscule amount of material by generating genome-wide overlapping amplicons, which has led to its success during the Zika virus outbreak investigation in Brazil [5, 6]. Using leftover RNA samples from a standard RT-PCR diagnosis, the genomic information of SARS-CoV-2 can be decoded in less than a week. The data presented here provide an insight into the genetic repertoire, origins and viral lineages of SARS-CoV-2 in Thailand. The information is particularly important given the multiple introduction events into the country and the local expansion of the Thai-specific SARS-CoV-2 lineages.

### Genomic surveillance of SARS-CoV-2 populations in Thailand

We sequenced 27 anonymized RT-qPCR positive samples from Ramathibodi Hospital in Bangkok during March 13-28, 2020 (Supplementary Table 1) [EC approval number: MURA2020/676]. The hospital acted as one of the major diagnostic hubs for COVID-19 in Bangkok during the study period. Enrichment and amplification steps were done according to the ARTIC Network protocol with ARTIC primer version 2 [7]. The libraries were prepared using KAPA HyperPrep and KAPA Library Amplification kits and subsequently sequenced using with a MiSeq Reagent Kit v2 according to the manufactures’ protocols. Variant calling was performed using the ncov2019-artic-nf pipeline (https://github.com/connor-lab/ncov2019-artic-nf). Consensus sequences were used to construct the maximum-likelihood and Bayesian phylogenetic trees with recommended representatives from various lineages worldwide utilizing IQ-TREE 2.0 and BEAST v1.10.4, respectively (Supplementary Table 2) [8-10]. Interestingly, Thailand appears to have had multiple introduction events of SARS-CoV-2 into the country, as evidenced by at least six separate clusters in the maximum-likelihood tree (Figure 1 and Supplementary Figure 1) [11]. Based on a Pangolin classification system (Database version 27 April 2020), they are grouped into A, B.1, B.1.5 and B.4 lineages (Figure 1 and Supplementary Figure 1) [9, 12]. Considering the origins and lineage branches, these SARS-CoV-2 lineages are likely to have recent ancestors outside Thailand. For example, six of the samples in the B.1 lineage in our collection are grouped tightly with virus samples commonly found in the United States of America and Europe and collected during the same period (March 2020) as visualized by Nextstrain Timetree (Supplementary Figure 2). The constructed Bayesian tree displayed the similar structure to that of the maximum-likelihood approach (Supplementary Figure 3).

**Figure 1.**
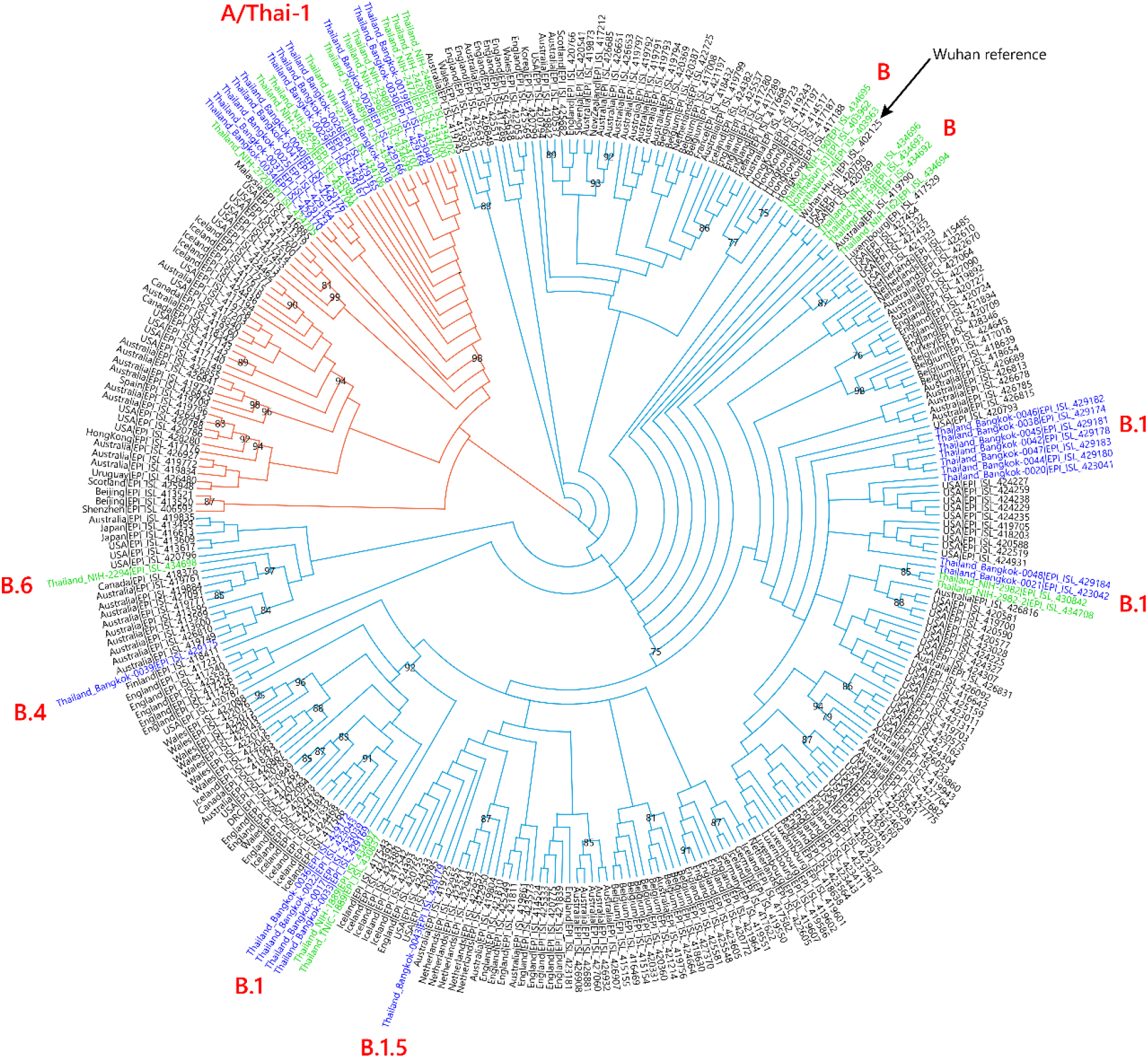
Cladogram based on a maximum-likelihood tree showing Thai populations of SARS-CoV-2. The cladogram represents a matching maximum-likelihood tree (1,000 bootstrap replicates) shown in Supplementary Figure 1. Genomes generated in this study are labelled in blue with remaining lineage representatives labelled in black. Bootstrap values (≥ 75) are shown at the nodes. Thai virus genomes independently generated by other groups and deposited in GISAID are labelled in green. Genomes collected from January 2020 are similar to that of reference Wuhan-Hu-1. Our data showed at least four independently lineages, and two additional events of lineage B from January 2020 and lineage B.6 (shown in green), could be recognized as potential introduction events. The A and B lineages are coloured in orange and cyan, respectively.

It is worth noting the local expansion of a putative Thai specific lineage (Figure 2). This cluster of viruses has passed the criteria of a novel lineage as follows: (a) exhibits two shared nucleotide differences from the ancestral lineage [cut point ≥ 1], (b) contains 11 genomes with > 95% of the genome sequenced [cut point ≥ 5], (c) exhibits one shared nucleotide change (27,877G→U) among the ongoing transmission groups from March 2020 [cut point ≥ 1], and (d) has 99% bootstrap value for the lineage defining node [cut point > 70%] [9]. This lineage, designated A/Thai-1 (Figure 1 and 2), descended from the original A lineage (based on the maximum-likelihood based classification system), which was first reported in China before expanding into various countries in Asia, Europe, North America, South America and Australia [9]. This A/Thai-1 branch is separated from the rest of the original A lineage and subgroups. Upon visual inspection in Nextstrain, only one Malaysian sample (MKAK-CL-2020-5096) is the closest to A/Thai-1, but only with 63% bootstrap value and one shared lineage-specific nucleotide substitution (4,390G→U) (Supplementary Figure 4). Non-synonymous mutations unique to A/Thai-1 are 20,134G→U (ORF1b) and 24,047G→A (Spike protein) (the full mutation list is shown in Table 1). Among the changes, 20,134G→U mutation has been independently found in two samples in lineage B.1 from the Netherlands and USA. It remains to be determined with a larger sample size whether this is the result of convergent evolution or genetic recombination. This pattern of homoplasy was also hypothetically linked to putative RNA editing [13].

**Figure 2.**
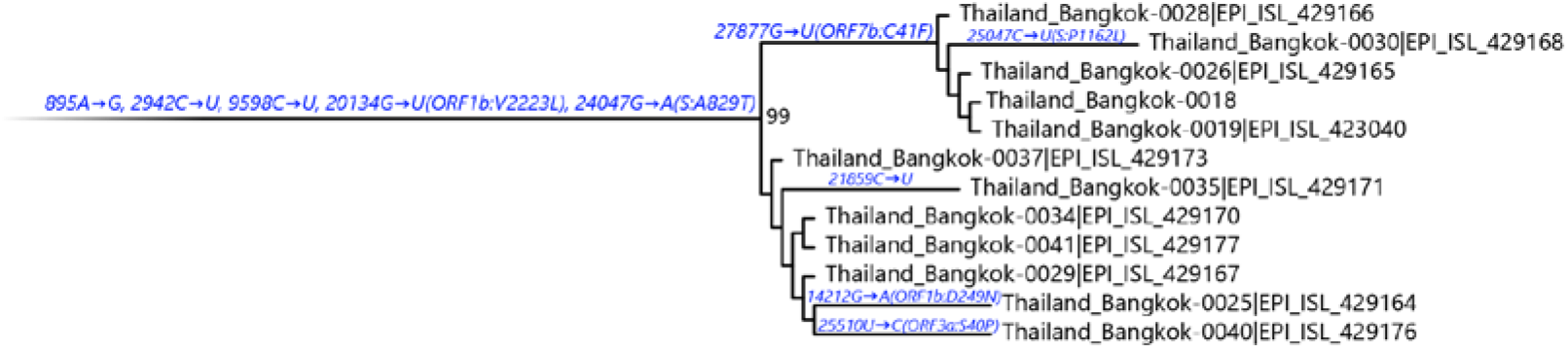
Maximum-likelihood tree containing A/Thai-1 subset. For visualization of A/Thai-1 lineage, the A/Thai-1 genomes from Figure 1 generated in this study are presented. The lineage defining node has a bootstrap value of 99 (1,000 replicates). Mutations that define each branch point are shown. 20,134G→U in Thailand_Bangkok-0030 could not be called.

**Table 1.** Mutations present in A/Thai-1 in comparison with Wuhan-Hu-1.

| Position | Ref | Alt | Syn/<br>Non-syn | Gene | Samples | Note |
| --- | --- | --- | --- | --- | --- | --- |
| 895 | A | G | S | ORF1a | 0018, 0019, 0025, 0026, 0028, 0029, 0030, 0034, 0035, 0037, 0040, 0041 | Unique to A/Thai-1 |
| 2942 | C | U | S | ORF1a | 0018, 0019, 0025, 0026, 0028, 0029, 0030, 0034, 0035, 0037, 0040, 0041 | Unique to A/Thai-1 |
| 4390 | G | U | NS | ORF1a | 0018, 0019, 0025, 0026, 0028, 0029, 0030, 0034, 0035, 0037, 0040, 0041 | Shared with Malaysia/MKAK-CL-2020-2096 (A lineage) |
| 8782 | C | U | S | ORF1a | 0018, 0019, 0025, 0026, 0028, 0029, 0030, 0034, 0035, 0037, 0040, 0041 | Shared among ancestral A lineages |
| 9598 | C | U | S | ORF1a | 0018, 0019, 0025, 0026, 0028, 0029, 0030, 0034, 0035, 0037, 0040, 0041 | Shared with Scotland/EDB2081 (B.1 lineage) |
| 14212 | G | A | NS | ORF1b | 0025 | - |
| 20134 | G | U | NS | ORF1b | 0018, 0019, 0025, 0026, 0028, 0029, 0034, 0035, 0037, 0040, 0041 | Cannot be called in 0030; Shared with Netherlands/NA_300 and USA/UN-UW-5172 (B.1 lineage) |
| 21859 | C | U | S | S | 0035 | - |
| 24047 | G | A | NS | S | 0018, 0019, 0025, 0026, 0028, 0029, 0030, 0034, 0035, 0037, 0040, 0041 | Unique to A/Thai-1 |
| 25047 | C | U | NS | S | 0030 | - |
| 25510 | U | C | NS | ORF3a | 0040 | - |
| 27694-27697 | TTT<br>C | - | Frame-shift<br>deletion | ORF7a | 0018 | - |
| 27877 | G | U | NS | ORF7b | 0018, 0019, 0026, 0028, 0030 | Shared with Taiwan/170 and USA/UT-00514 (B.1 lineage) |
| 28144 | U | C | NS | ORF8 | 0018, 0019, 0025, 0026, 0028, 0029, 0030, 0034, 0035, 0037, 0040, 0041 | Shared among ancestral A lineages |

Thailand/Bangkok-0018, a sample in the A/Thai-1 lineage, contains a 4-nt frame-shift deletion at position 27,694-27,697, causing a premature truncation in ORF7a, which now contains five altered amino acid residues and loses the 16 original C-terminal residues (Figure 3). The deletion was confirmed by Sanger sequencing twice using two independent RT-PCR reactions (Supplementary Figure 5). The frame-shift mutation alters approximately one-sixth (21/121 residues) of the ORF7a protein. Based on protein homology to SARS-CoV, the missing region corresponds to a transmembrane helix and an ER retrieval motif, required for antagonizing a host antiviral factor [14, 15]. One sample from Arizona, USA also contains an 81-nt in-frame deletion in the ORF7a gene [16]. So far, only one sample in A/Thai-1 appears to have this frame-shift deletion. It is tempting to speculate on the relationship between ORF7a deletions and virus attenuation. However, further investigations by laboratory-based functional experiments are needed before reaching any conclusion on their biological and clinical significance.

**Figure 3.**
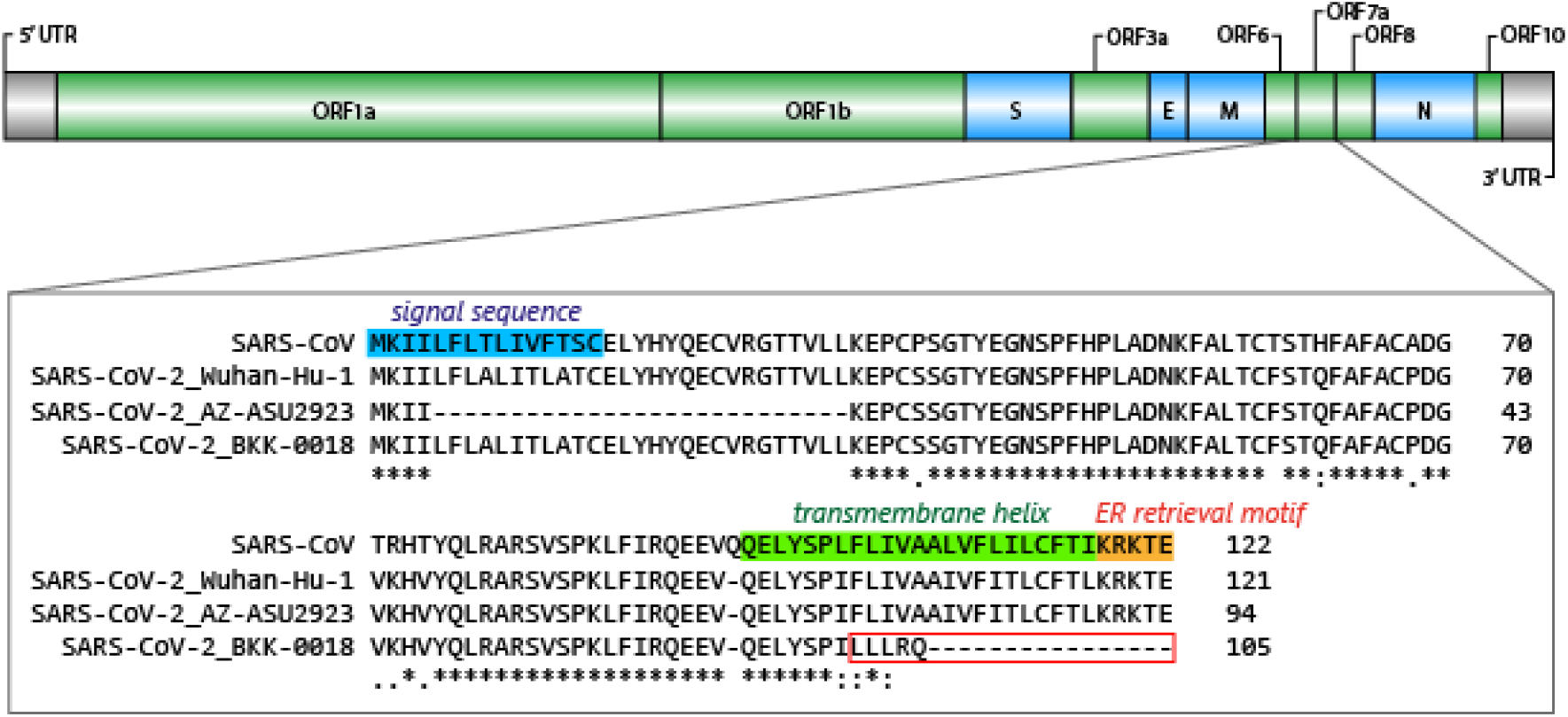
Diagram depicting Thailand/Bangkok-0018 frame-shift deletion in ORF7a. The upper diagram shows the gene organization of the SARS-CoV-2 genome. The functional domains of ORF7a are indicated based on SARS-CoV. Sequences from SARS-CoV, the SARS-CoV-2 Wuhan-Hu-1 reference, the Arizona sample with an 81-nucleotide deletion (EPI_ISL_424669) and Thailand/Bangkok-0018 are aligned to demonstrate the altered region in red box.

When the analysis was extended to 22 additional genomes, independently deposited in GISAID by the Thai National Influenza Center and the Thai National Institute of Health, samples collected in January 2020 are grouped closely with the B lineage from China including the Wuhan-Hu-1 reference (Supplementary Table 3). The genetic repertoires from this additional collection also support the notion of multiple virus lineages introduced into Thailand. Nine of them also fall into A/Thai-1, making it the largest lineage in Thailand during the period of March 2020 (22/49 genomes).

### Implication of the findings

Genomic surveillance is likely to be pivotal in the identification and the elimination of transmission cohorts and chains [17, 18]. The genetic composition presented here suggests the necessity for screening and monitoring international travelers during the period of COVID-19 pandemic. The local expansion of A/Thai-1 has created a new evolutionary branch unique to Thailand, which inevitably requires this lineage to be investigated for its compatibility to diagnosis and vaccine tools under development.

## Data Availability

The data that support the findings of this study are openly available at https://www.gisaid.org/ as described in the manuscript.

## Acknowledgment

The work here was supported by Ramathibodi Foundation, TCELS, NRCT and Mahidol University. The authors acknowledge NSTDA Supercomputer Center (ThaiSC) for providing computing resources for this work. We are grateful for the comment and suggestion from P. Wilairat and Y. Yuthavong. We appreciate the contribution from K. Joonlasak, A. Huang, A.R. Jones, S. Fernandez and C. Klungthong (Armed Forces Research Institute of Medical Sciences). The computational aspects of this research were supported by the Wellcome Trust Core Award Grant Number 203141/Z/16/Z and the NIHR Oxford BRC. The views expressed are those of the author(s) and not necessarily those of the NHS, the NIHR or the Department of Health.

---

**Supplementary Figure 1.**
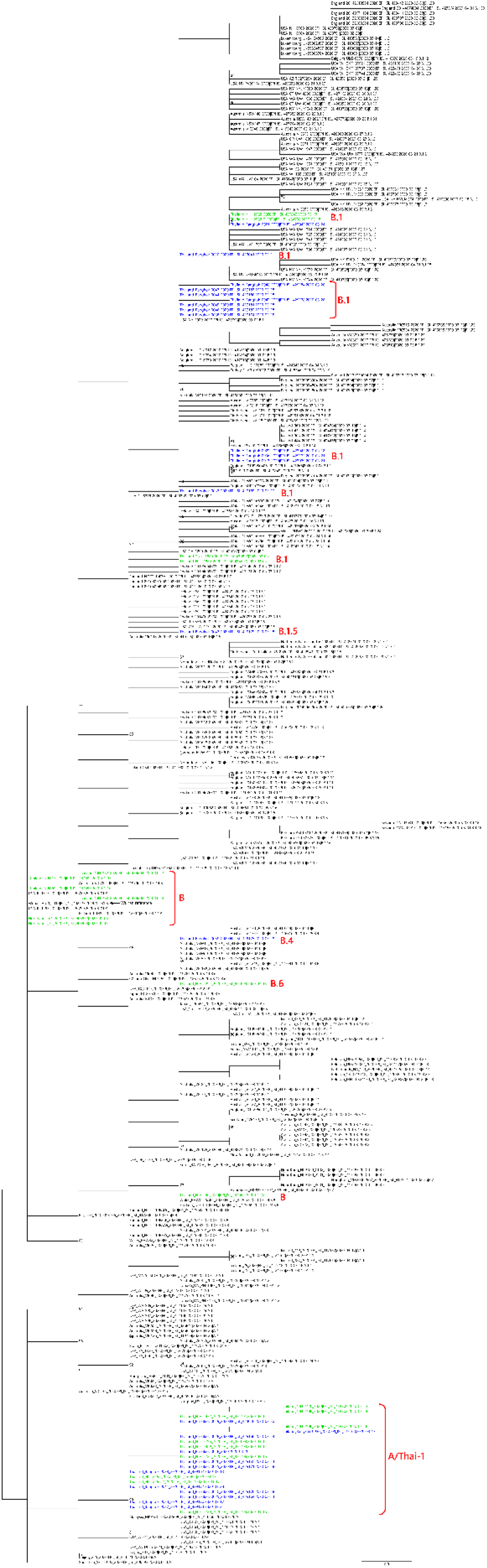
Maximum-likelihood tree used for the cladogram depiction in Figure 1. The labels and colors are similar to those in Figure 1.

**Supplementary Figure 2.**
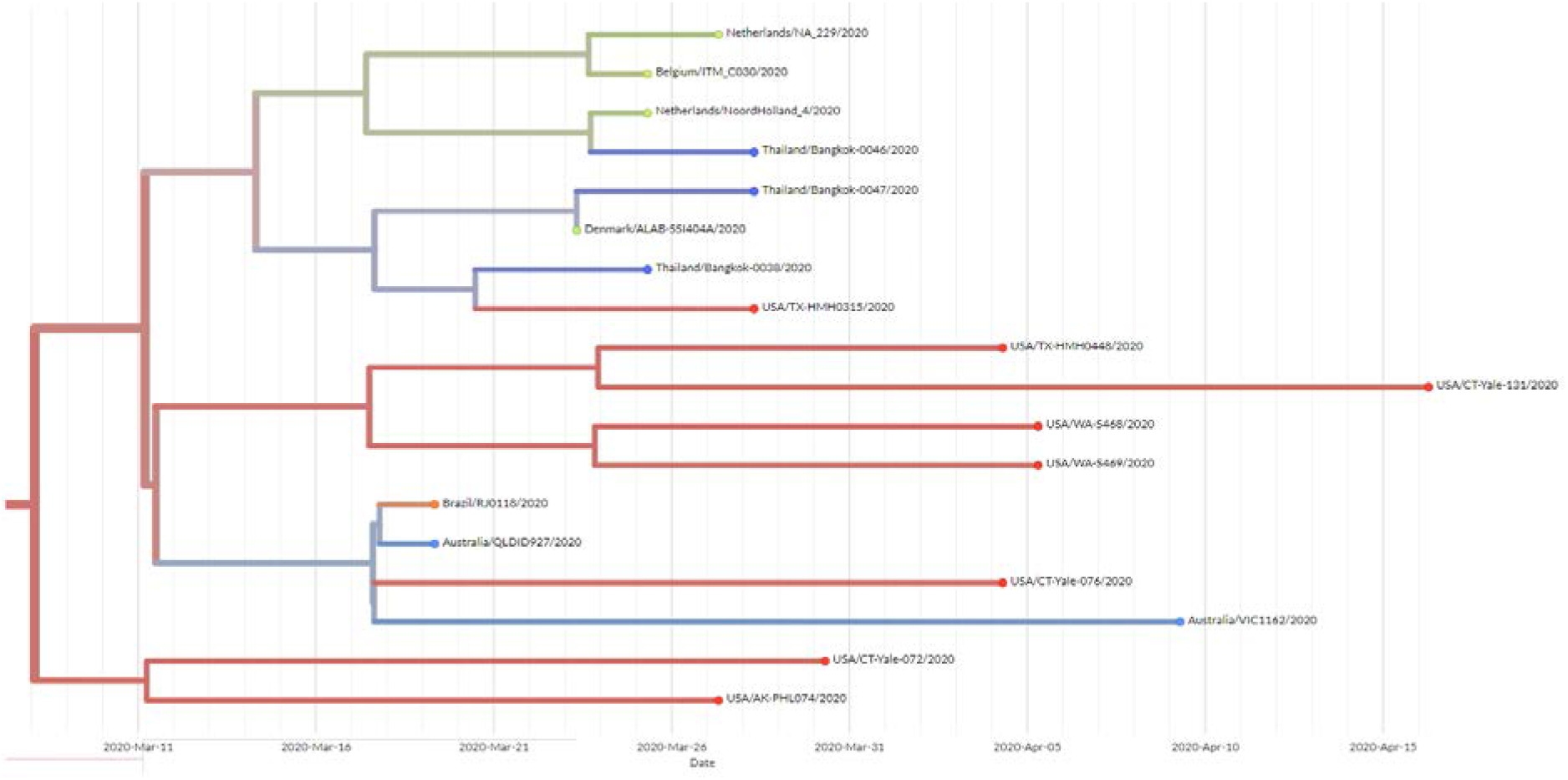
Nextstrain Timetree presenting the grouping of the Thai SARS-CoV-2 B1 samples with the virus genomes from other countries during the same period. The data was retrieved on 6 May 2020.

**Supplementary Figure 3.**
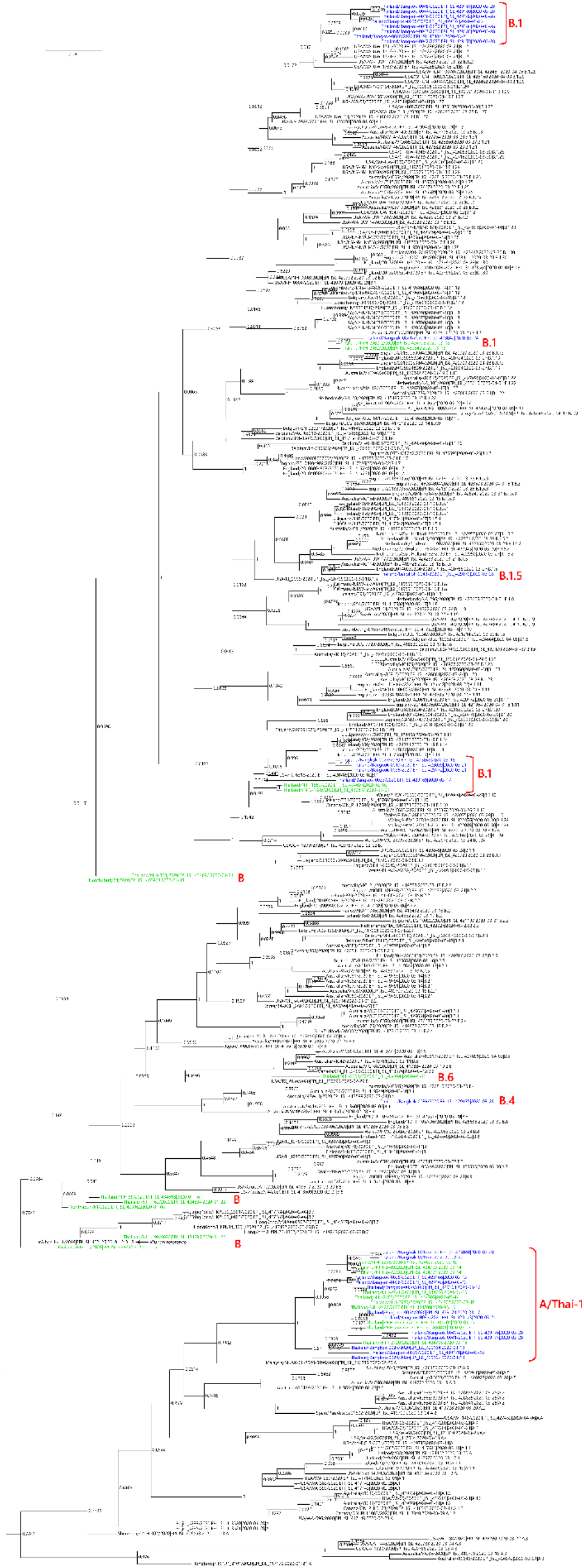
Bayesian phylogenetic tree of the SARS-CoV-2. Tree plotting was done with the same sample sets used for the maximum-likelihood tree.

**Supplementary Figure 4.**
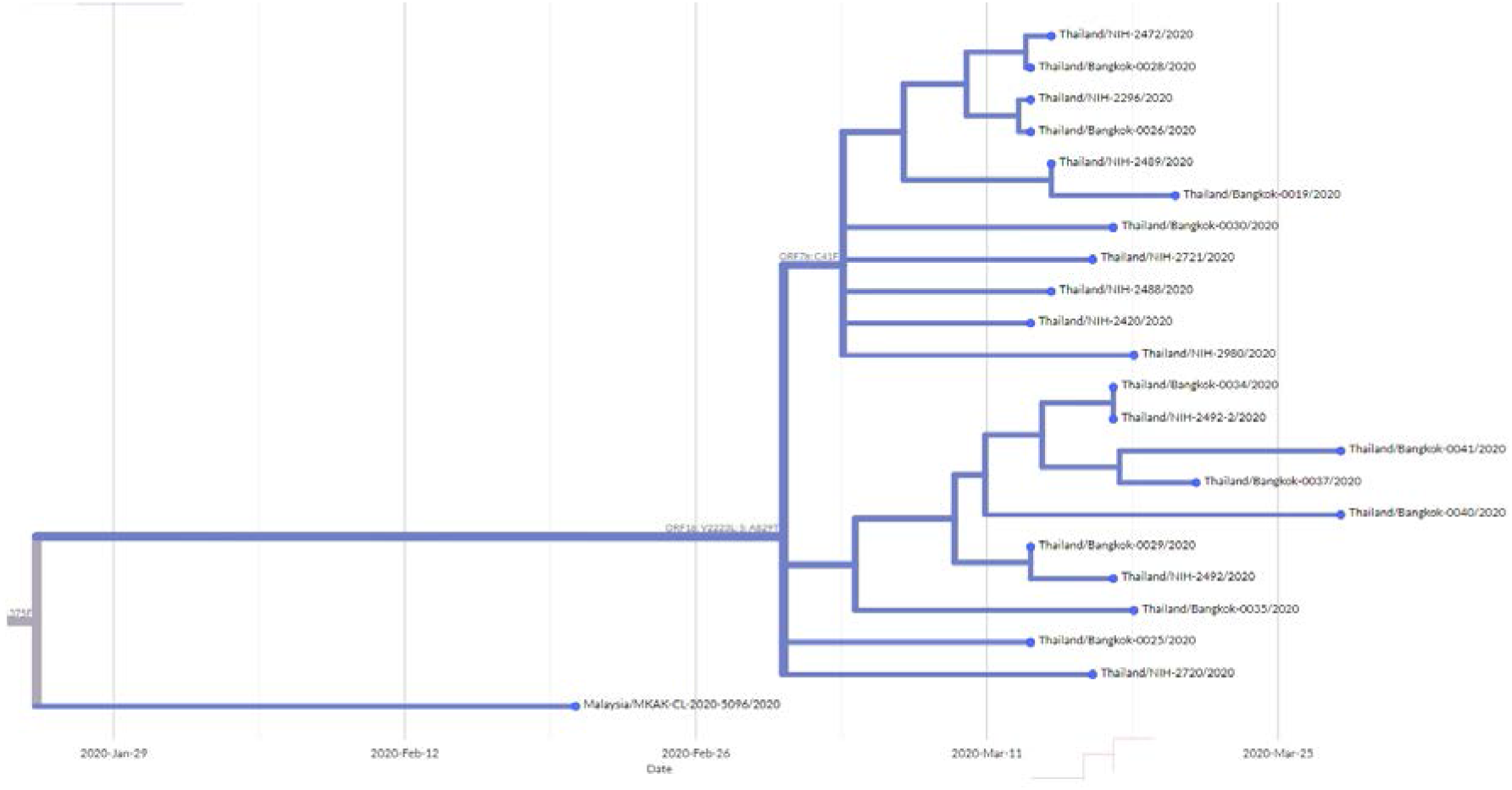
Nextstrain Timetree representing the A/Thai-1 lineage using the data from GISAID. The Malaysian virus genome MKAK-CL-2020-5096 is the closest one in the tree. The data was retrieved on 6 May 2020.

**Supplementary Figure 5.**
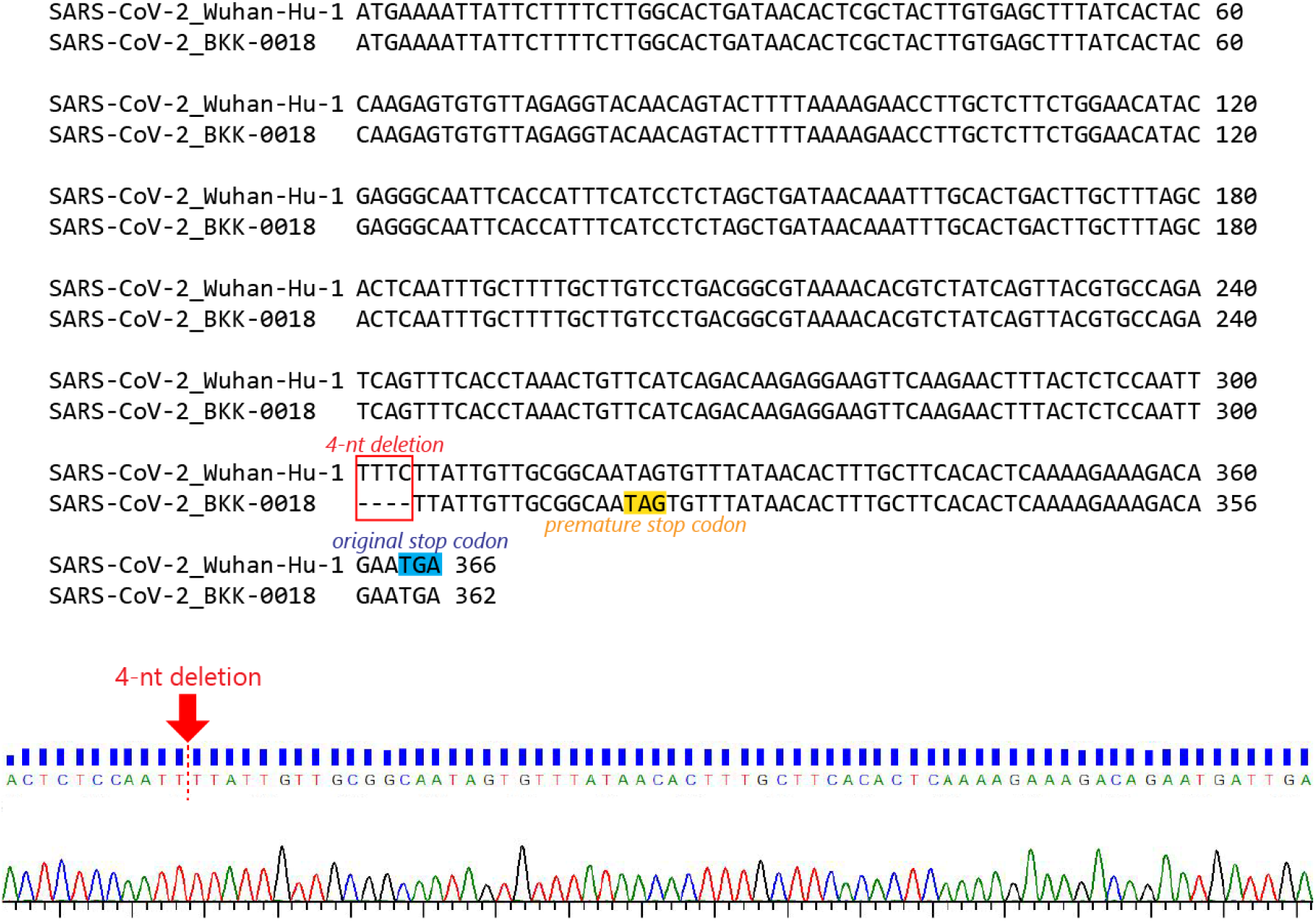
Sanger sequencing result confirming the ORF7a deletion in BKK-0018. A sequencing chromatogram from the DNA regions (upper panel) corresponding to the deletion site (red arrow) is shown. The 4-nt deletion site marked in the red box causes a premature stop codon.

**Supplementary Table 1:** List of samples used for genome.

| **Name** | **GISAID ID** | **Collection date** |
| --- | --- | --- |
| Thailand/Bangkok-0017/2020 | EPI_ISL_423039 | 2020-03-21 |
| Thailand/Bangkok-0018/2020 | EPI_ISL_450723 | 2020-03-20 |
| Thailand/Bangkok-0019/2020 | EPI_ISL_423040 | 2020-03-20 |
| Thailand/Bangkok-0020/2020 | EPI_ISL_423041 | 2020-03-21 |
| Thailand/Bangkok-0021/2020 | EPI_ISL_423042 | 2020-03-24 |
| Thailand/Bangkok-0022/2020 | EPI_ISL_423043 | 2020-03-18 |
| Thailand/Bangkok-0025/2020 | EPI_ISL_429164 | 2020-03-13 |
| Thailand/Bangkok-0026/2020 | EPI_ISL_429165 | 2020-03-13 |
| Thailand/Bangkok-0028/2020 | EPI_ISL_429166 | 2020-03-13 |
| Thailand/Bangkok-0029/2020 | EPI_ISL_429167 | 2020-03-13 |
| Thailand/Bangkok-0030/2020 | EPI_ISL_429168 | 2020-03-17 |
| Thailand/Bangkok-0033/2020 | EPI_ISL_429169 | 2020-03-17 |
| Thailand/Bangkok-0034/2020 | EPI_ISL_429170 | 2020-03-17 |
| Thailand/Bangkok-0035/2020 | EPI_ISL_429171 | 2020-03-18 |
| Thailand/Bangkok-0036/2020 | EPI_ISL_429172 | 2020-03-21 |
| Thailand/Bangkok-0037/2020 | EPI_ISL_429173 | 2020-03-21 |
| Thailand/Bangkok-0038/2020 | EPI_ISL_429174 | 2020-03-25 |
| Thailand/Bangkok-0039/2020 | EPI_ISL_429175 | 2020-03-28 |
| Thailand/Bangkok-0040/2020 | EPI_ISL_429176 | 2020-03-28 |
| Thailand/Bangkok-0041/2020 | EPI_ISL_429177 | 2020-03-28 |
| Thailand/Bangkok-0042/2020 | EPI_ISL_429178 | 2020-03-28 |
| Thailand/Bangkok-0043/2020 | EPI_ISL_429179 | 2020-03-28 |
| Thailand/Bangkok-0044/2020 | EPI_ISL_429180 | 2020-03-28 |
| Thailand/Bangkok-0045/2020 | EPI_ISL_429181 | 2020-03-28 |
| Thailand/Bangkok-0046/2020 | EPI_ISL_429182 | 2020-03-28 |
| Thailand/Bangkok-0047/2020 | EPI_ISL_429183 | 2020-03-28 |
| Thailand/Bangkok-0048/2020 | EPI_ISL_429184 | 2020-03-28 |

**Supplementary Table 2:**
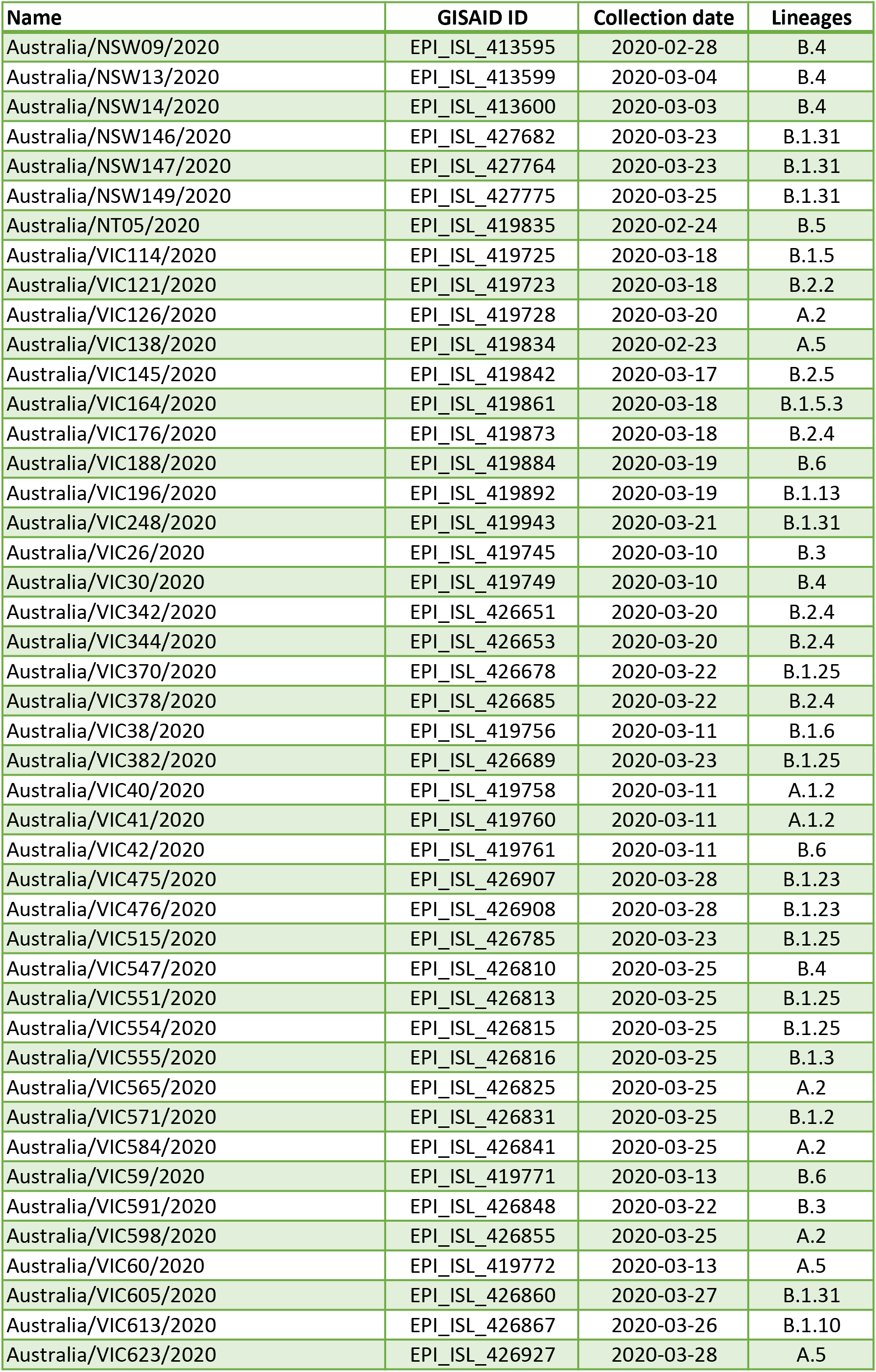
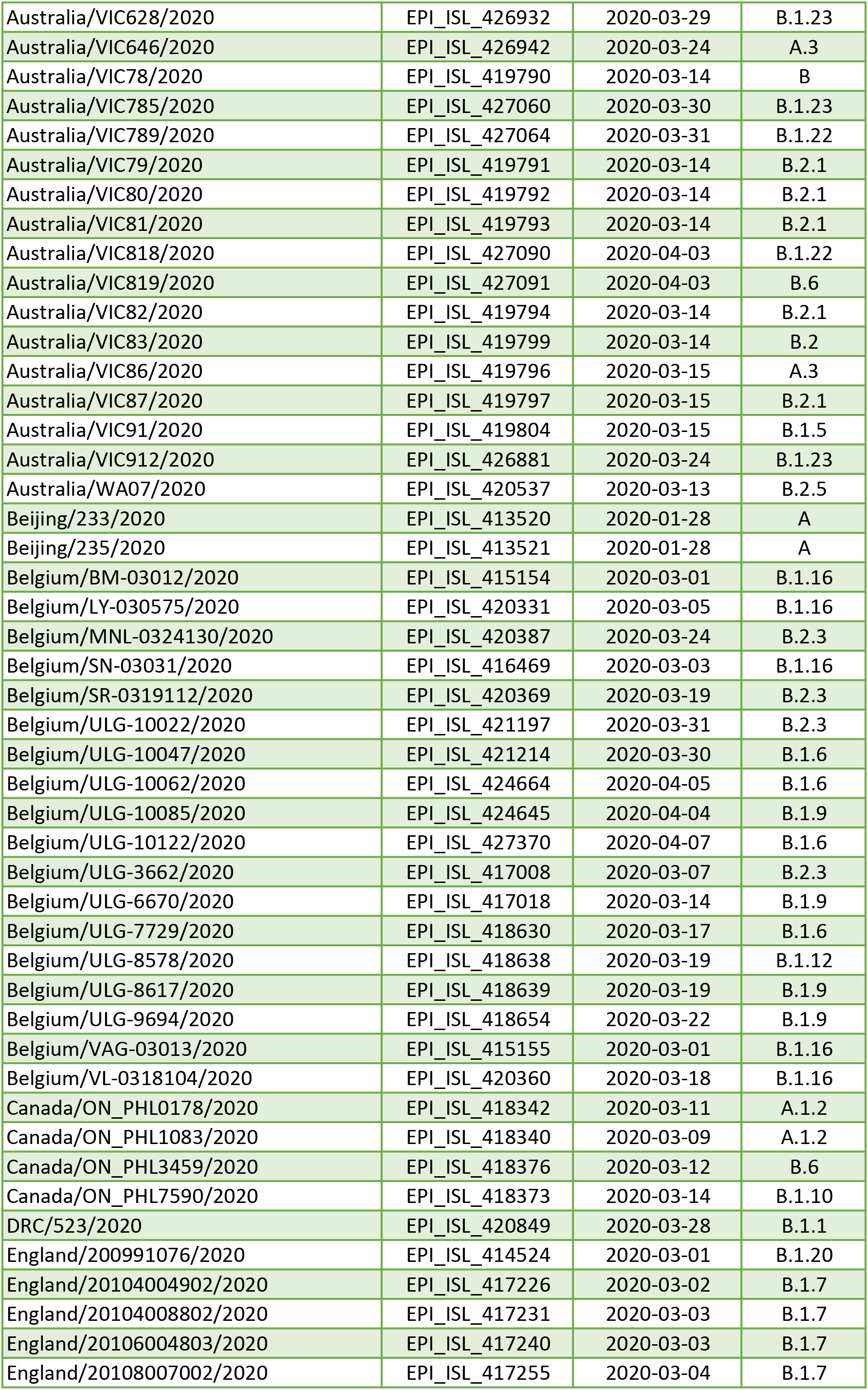
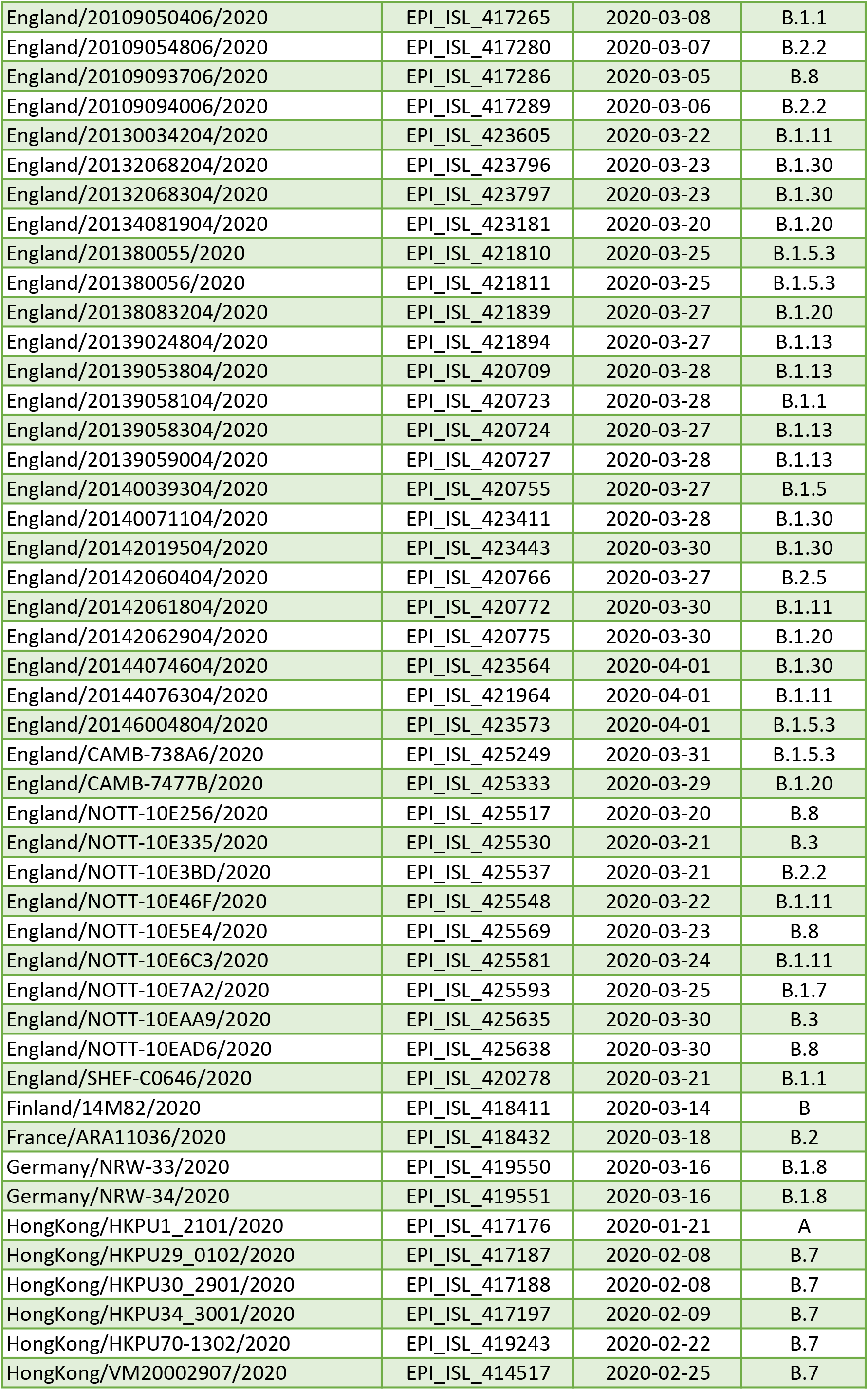
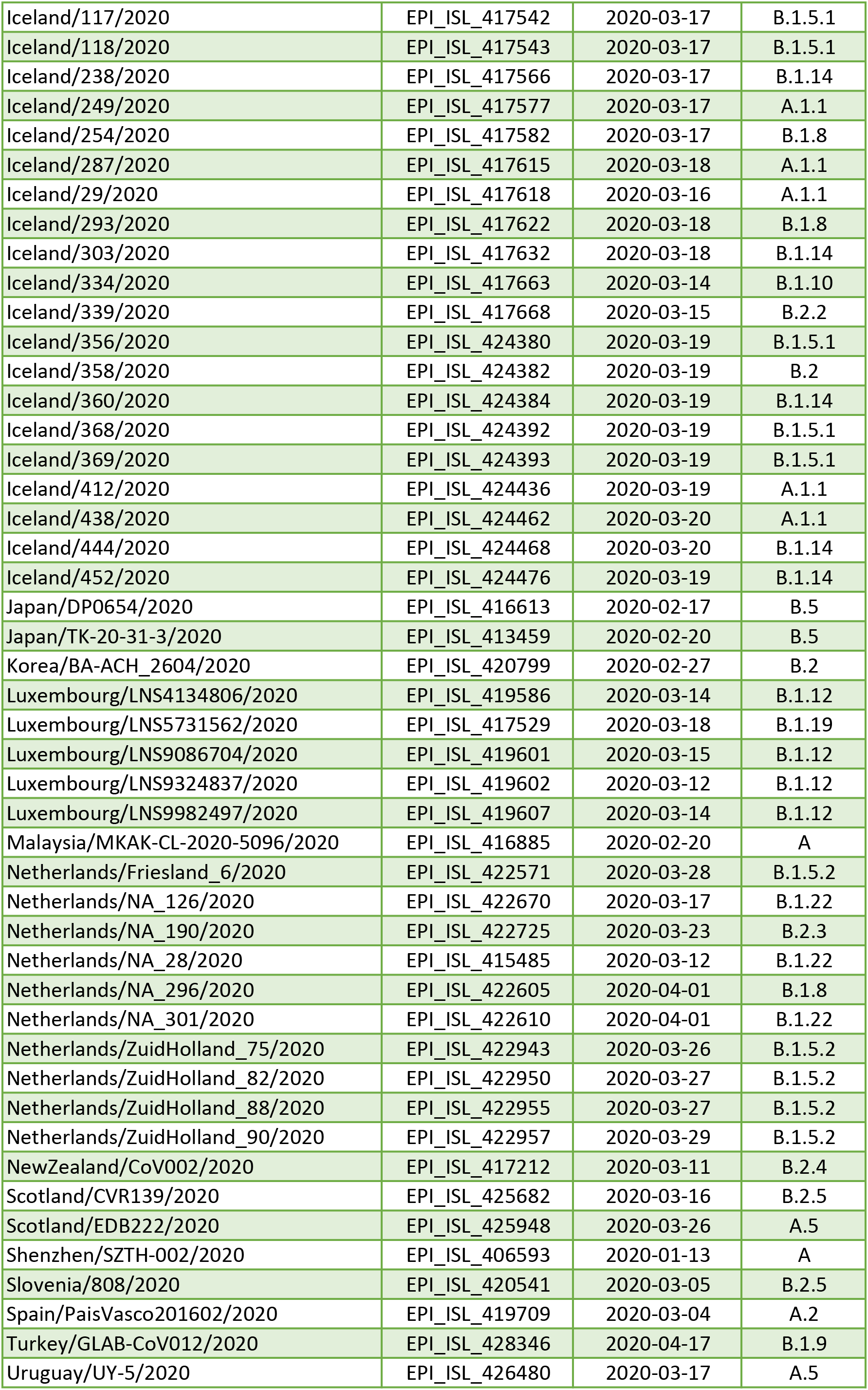
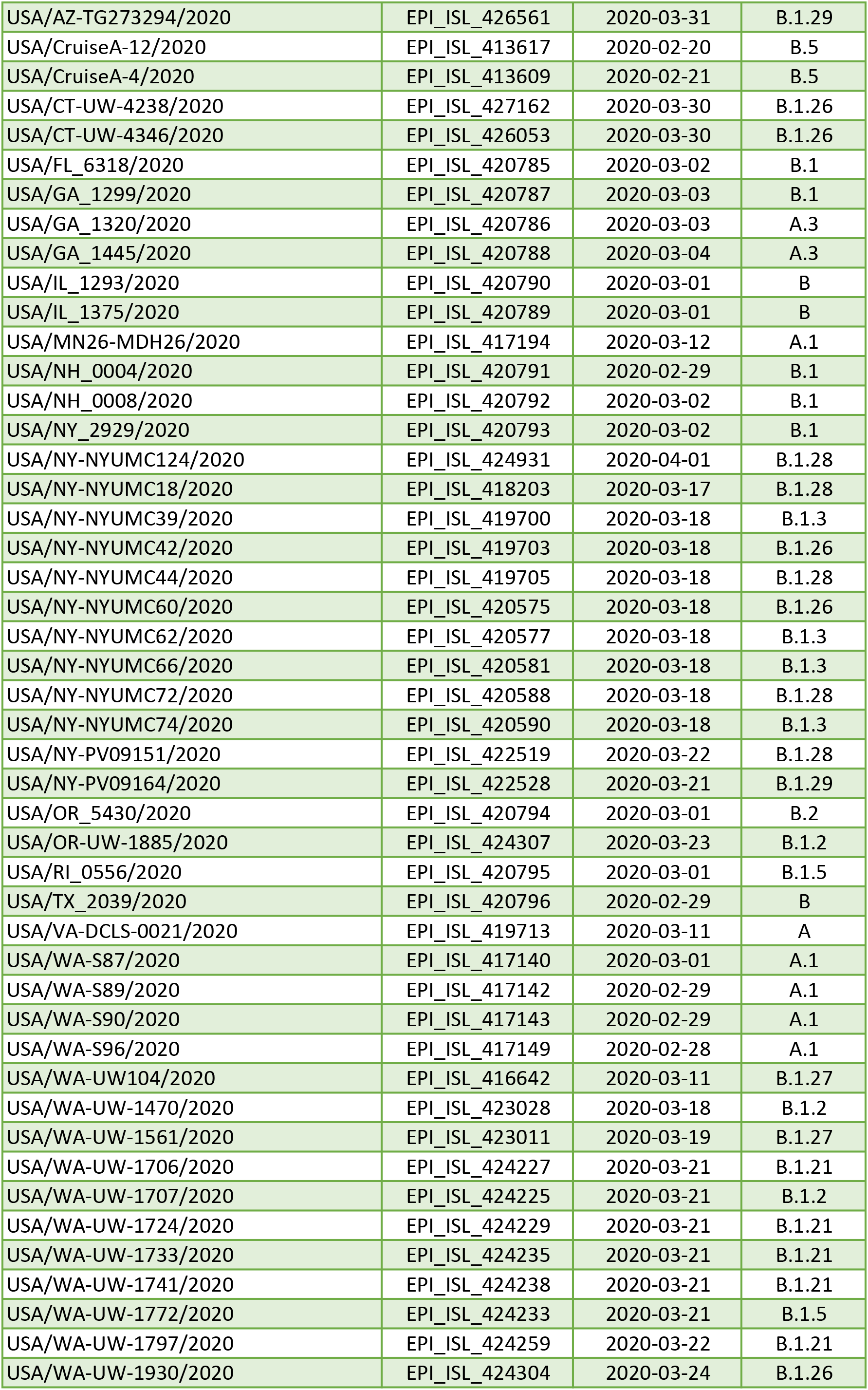
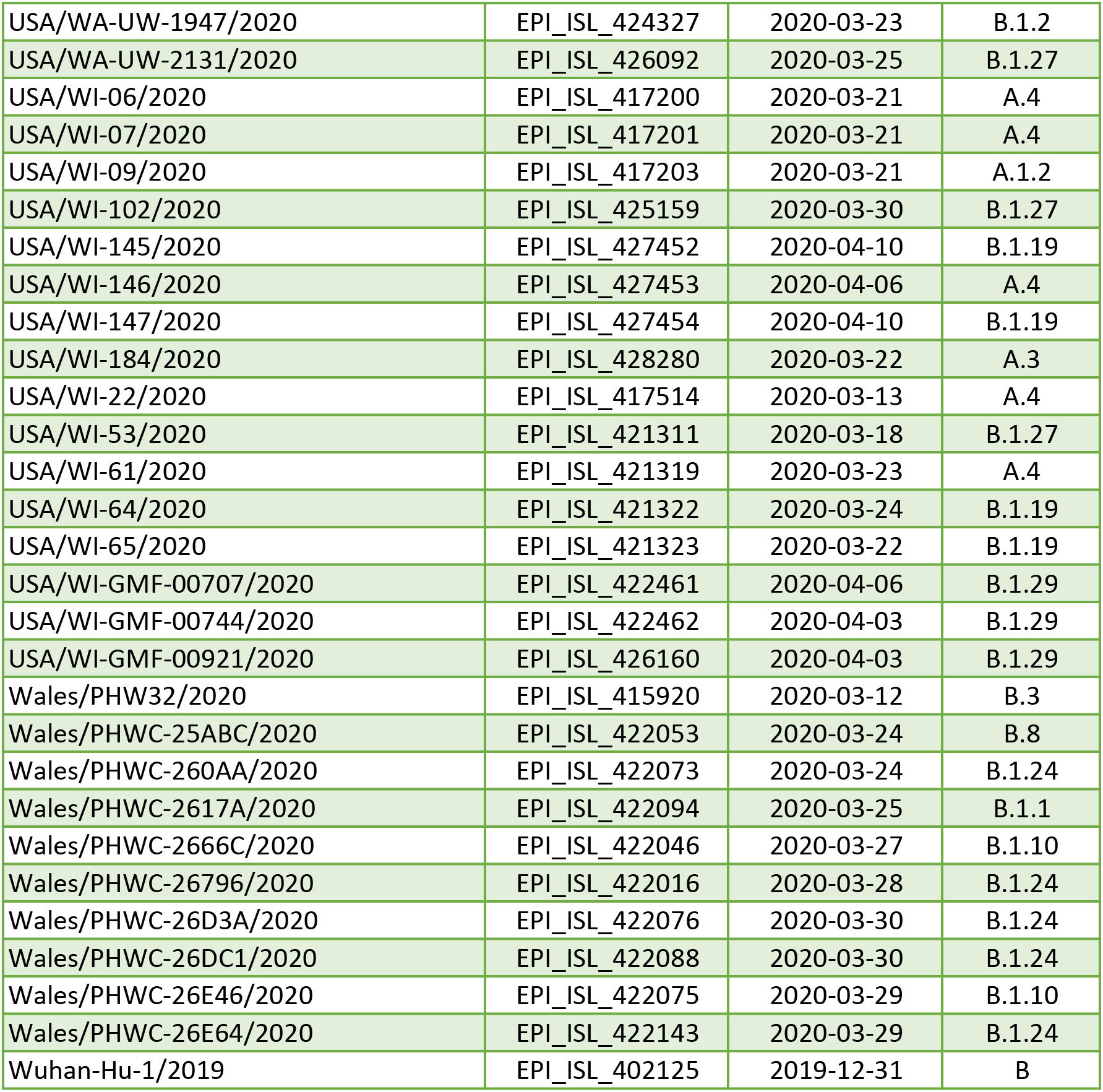
List of representatives used for phylogenetics analyses.

**Supplementary Table 3:** genomes from the Thai samples deposited in GISAID by other groups.

| Name | GISAID ID | Collection date |
| --- | --- | --- |
| Thailand/NIH-15/2020 | EPI_ISL_434692 | 2020-01-05 |
| Thailand/NIH-162/2020 | EPI_ISL_434694 | 2020-01-22 |
| Thailand/NIH-1889/2020 | EPI_ISL_434697 | 2020-03-03 |
| Thailand/NIH-190/2020 | EPI_ISL_434695 | 2020-01-22 |
| Thailand/NIH-2294/2020 | EPI_ISL_434698 | 2020-03-13 |
| Thailand/NIH-2296/2020 | EPI_ISL_434699 | 2020-03-13 |
| Thailand/NIH-2420/2020 | EPI_ISL_434700 | 2020-03-13 |
| Thailand/NIH-2472/2020 | EPI_ISL_434701 | 2020-03-14 |
| Thailand/NIH-2488/2020 | EPI_ISL_434702 | 2020-03-14 |
| Thailand/NIH-2489/2020 | EPI_ISL_434703 | 2020-03-14 |
| Thailand/NIH-2492/2020 | EPI_ISL_430841 | 2020-03-17 |
| Thailand/NIH-2492-2/2020 | EPI_ISL_434704 | 2020-03-17 |
| Thailand/NIH-2720/2020 | EPI_ISL_434705 | 2020-03-16 |
| Thailand/NIH-2721/2020 | EPI_ISL_434706 | 2020-03-16 |
| Thailand/NIH-2980/2020 | EPI_ISL_434707 | 2020-03-18 |
| Thailand/NIH-2982/2020 | EPI_ISL_430842 | 2020-03-18 |
| Thailand/NIH-2982-2/2020 | EPI_ISL_434708 | 2020-03-18 |
| Thailand/NIH-363/2020 | EPI_ISL_434696 | 2020-01-24 |
| Thailand/NIH-59/2020 | EPI_ISL_434693 | 2020-01-16 |
| Thailand/TNIC-1889/2020 | EPI_ISL_430837 | 2020-03-03 |

